# Machine learning-based calculation of neurovascular compression surface area correlates with post-microvascular decompression pain outcomes for trigeminal neuralgia

**DOI:** 10.1101/2025.03.25.25324660

**Authors:** Xihang Wang, Kyra M. Halbert-Elliot, Michael E. Xie, Oishika Das, Kathleen R. Ran, Bryan Dong, Mostafa Abdulrahim, Christopher M. Jackson, Michael Lim, Judy Huang, Vivek S. Yedavalli, Chetan Bettegowda, Risheng Xu

## Abstract

**Background:** Machine learning-generated segmentations of the trigeminal nerve and nearby blood vessels have the potential to quantify the magnitude of neurovascular compression (NVC) in patients with trigeminal neuralgia (TN). This study applies the nnU-Net machine learning method to create segmentations of the trigeminal nerve region from patient MRIs and correlate resulting quantitative NVC metrics with postoperative TN outcomes.

**Methods:** MRIs from patients undergoing microvascular decompression (MVD) for TN from 2019 to 2022 at a single tertiary care facility were split into training, testing, and inference datasets. The trigeminal nerve and surrounding vasculature were manually labeled (i.e., segmented) in the training and testing datasets to create ground truth (GT) segmentations. nnU-Net was trained on GT segmentations in the training dataset, and predicted segmentations were evaluated using the testing dataset via the F1 score, IoU score, and paired comparison of resulting metrics. To contextualize nnU-Net performance, a manual SE-ResNet152 model was trained and deployed using the same datasets. Predicted nnU-Net segmentations in the inference dataset were then correlated with the rate of post-MVD pain recurrence.

**Results:** Of 366 total GT segmentations, 302 (82.5%) trained the nnU-Net model and 64 (17.5%) validated the predicted segmentations. The nnU-Net model’s F1 and IoU scores on the testing dataset were 0.797±0.011 and 0.714±0.011, respectively, which were higher than those for SE-ResNet152. The sensitivity and specificity of nnU-Net’s ability to detect NVC were 91.3% and 66.7%, respectively. Surface area of NVC calculated from nnU-Net and GT segmentations were statistically similar. Deployed on the inference dataset (n=100), higher surface area of NVC was observed in patients without pain recurrence following MVD than patients with pain recurrence (*p=0*.*008*). Finally, higher surface area of NVC (hazards ratio [HR] 0.914 per mm^2^, 95% confidence interval [CI] 0.848–0.985, *p=0*.*019*) and presence of NVC (HR 0.369 relative to absent NVC, 95% CI 0.156–0.876, *p=0*.*024*) were both associated with a significantly decreased risk of pain recurrence in Cox proportional hazards models.

**Conclusion:** nnU-Net can generate high-fidelity segmentations of the trigeminal nerve region, and the resulting NVC surface area metric is significantly associated with post-MVD pain recurrence. nnU-Net can be a standardized tool to evaluate NVC severity for patients seeking TN treatment.

## Introduction

Trigeminal neuralgia (TN) is a neuropathic condition characterized by severe unilateral facial pain following one or more branch distributions of the trigeminal nerve.^1^ Though the exact mechanisms underlying TN are still under investigation, one common hypothesis is that TN arises in part from compression of the trigeminal nerve by nearby arteries and veins in the cerebrospinal fluid space.^1,2^ This neurovascular contact (NVC) subsequently wears down the myelin sheath surrounding the trigeminal nerve, leading to paroxysmal nerve firing and the perception of intense facial pain.^1,2^ In rare cases, other compressing structures such as arteriovenous malformations and tumors may drive TN symptomatology through a similar mechanism.^2^

Assessment of the trigeminal nerve region on brain magnetic resonance imaging (MRI) is key in the preoperative evaluation for TN treatment, particularly in determining which surgical option to pursue and the likelihood of operative success. If clear contact between the affected trigeminal nerve and nearby vessels is identified, microvascular decompression (MVD), in which the offending vessel is separated from the nerve, is sensible. Given MVD is the most invasive of TN treatments, confidence in outcomes can aid in counseling and shared decision making. The magnitude of NVC has been associated with clinical features of TN, such as pain outcomes following MVD.^3,4^ However, it is difficult to quantify the severity of NVC with direct MRI reads as they lack objective measurements for nerve-vessel contact and are therefore subject to inter-reader variability.^5–7^ Segmentations—labeled three-dimensional (3D) reconstructions of the trigeminal nerve and surrounding blood vessels derived from preoperative MRI scans—can provide more precise metrics that quantify the degree of NVC present to better characterize TN clinical outcomes.^4,8–12^

In this study, we use deep learning methods to automatically generate segmentations of the trigeminal nerve and surrounding vasculature, from which quantitative metrics of NVC can be derived and correlated with clinical postoperative TN outcomes.^8,13–15^ Its near-instantaneous timescale and automated nature make deep learning-based segmentation an appealing alternative to traditional options, such as manual segmentation. To this end, our prior work evaluated the ability of U-Net based machine learning architectures to create 3D reconstructions of the trigeminal nerve and nearby vasculature.^15^ While the highest-performing architecture, SE-ResNet, produced reliable results from which geometric features could be extracted, its development process required the addition of a custom loss function and quantitative hyperparameter tuning. Such efforts limit the accessibility of a U-Net based model, especially if application to external datasets is desired. To address this barrier, we chose to evaluate the performance of nnU-Net at the same task. nnU-Net is a self-configuring deep learning pipeline for deploying U-Net on custom biomedical image segmentation tasks, comprised of automated hyperparameter optimization, architecture selection, and model training.^14^ The resulting nnU-Net-generated segmentations were compared with corresponding expert manual segmentations and SE-ResNet segmentations to evaluate their accuracy and performance before being correlated with postoperative TN pain outcomes.

## Methods

All patients ≥18 years of age who underwent MVD for TN from January 2019 to December 2022 at a single tertiary care facility were identified (IRB00338945). Exclusion criteria included prior MVD intervention, unavailable preoperative high-resolution 3-Tesla Constructive Interference in Steady State (CISS) or Fast Imaging Employing Steady-state Acquisition (FIESTA) brain MRI sequences, and image quality or metadata issues.^16,17^

Patients were split by MVD year into training, testing, and inference datasets. MRIs from patients who underwent MVD in 2020-2022 were randomly assigned into the training and testing datasets. Patients in the testing dataset would each have an nnU-Net-generated segmentation as well as a corresponding manually-created segmentation to serve as the gold standard (termed the “ground truth,” GT). Accordingly, paired comparison of the two was used to validate the accuracy of nnU-Net-generated segmentations. MRIs from patients who underwent MVD in 2019 were set aside to compose a live inference dataset to deploy the trained model and correlate results with clinical TN outcomes. This live inference dataset consisted of patients for whom nnU-Net would generate a predicted segmentation that was subsequently used to analyze post-MVD pain recurrence rates, thus evaluating the ability of nnU-Net to generate clinically-informative data. Of note, patients in this dataset do not have a corresponding GT segmentation, and patients with multiple sclerosis were excluded from this dataset because altered TN pathophysiology may bias results from correlation with clinical post-MVD outcomes.^18^ Each patient MRI contributed two segmentations, one trigeminal nerve on each side. Both segmentations from a single patient were assigned to the same dataset, thus maintaining independence in model training, testing, and inference.

### MRI pre-processing

The MRI pre-processing pipeline was completed in accordance with our prior work (**Figure 1**).^15^ The centroids of the bilateral trigeminal nerves on all MRI scans were identified by trained researchers and supervised by an attending neurosurgeon. A 30 mm by 30 mm by 30 mm volume was cropped around each centroid, and each voxel in the volume was rescaled to a size of 0.47 mm by 0.47 mm by 0.47 mm (64 voxels by 64 voxels by 64 voxels). The intensity values of the voxels were Z score-normalized.

**Figure 1.**
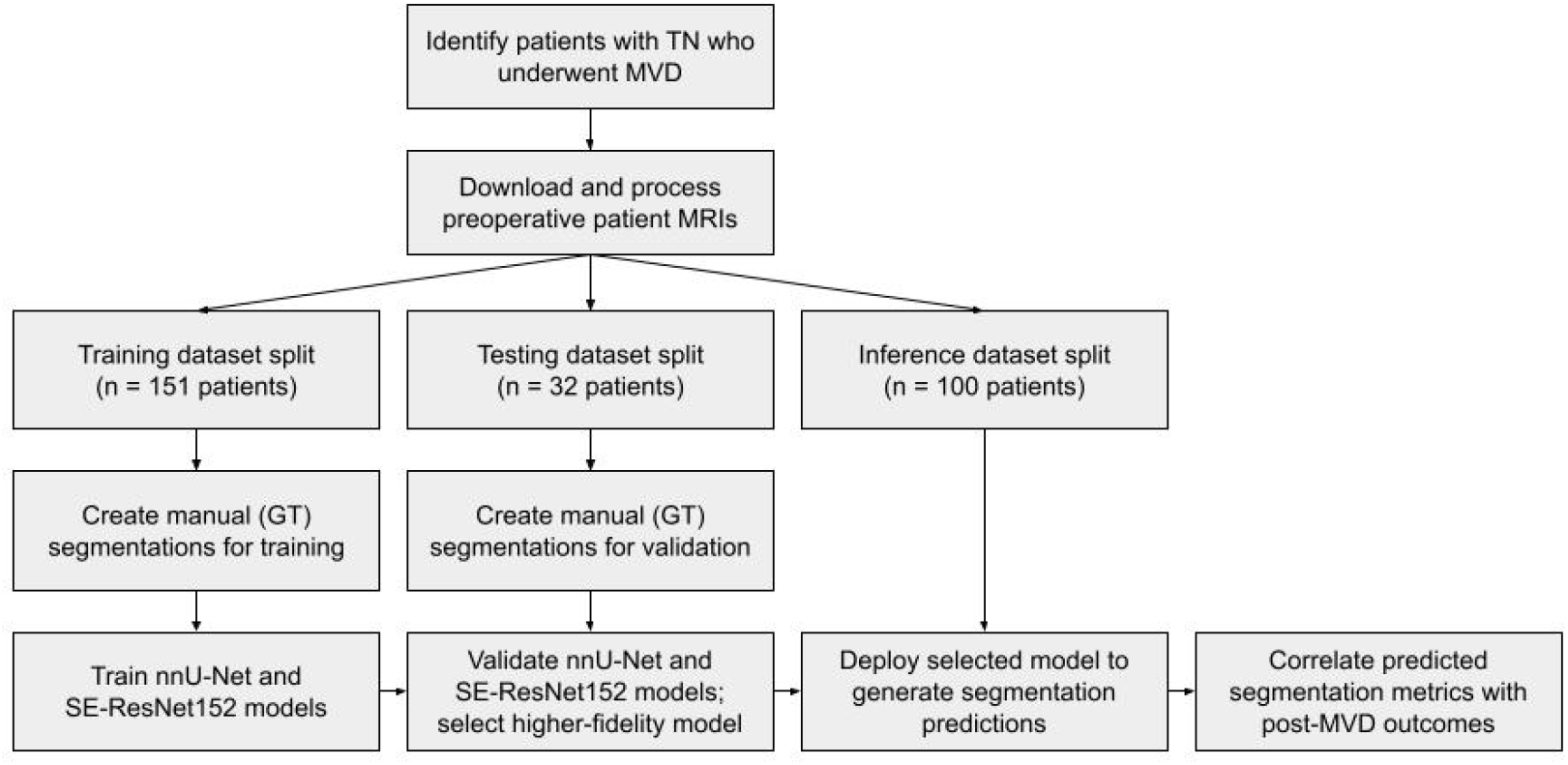
Schematic for the application of convolutional neural networks towards trigeminal nerve segmentation and subsequent correlation with TN clinical outcomes.

### Image segmentation

For all volumes in the training and testing datasets, each voxel was manually labeled by a trained researcher as trigeminal nerve, blood vessel, or background. These resulting manual segmentations comprised the ground truth (GT) dataset upon which the nnU-Net model was trained and validated. GT segmentations were cross-verified by multiple researchers, a board-certified neurosurgeon, and a board-certified neuroradiologist.

### Model training and testing

The GT training MRI volumes were loaded into the nnU-Net pipeline. The 3D full-resolution training configuration was selected for our model. The nnU-Net model was trained over five-fold cross-validation, undergoing 1000 epochs per fold. Model training was performed on the Johns Hopkins Joint High Performance Computing Exchange with GPU partition. Following training, the MRI volumes from the testing dataset were passed into the trained nnU-Net model, which then generated a segmentation for each testing MRI volume (**Figure 2, Figure 3**). This process was run from the ensemble of all five training folds. For comparison with nnU-Net, a U-Net model with an SE-ResNet backbone (SE-ResNet152) was trained on the same training dataset. The testing MRIs were passed through the trained model, yielding a predicted segmentation for each MRI from the SE-ResNet152 model that could be directly compared to segmentations produced from the nnU-Net model.

**Figure 2.**
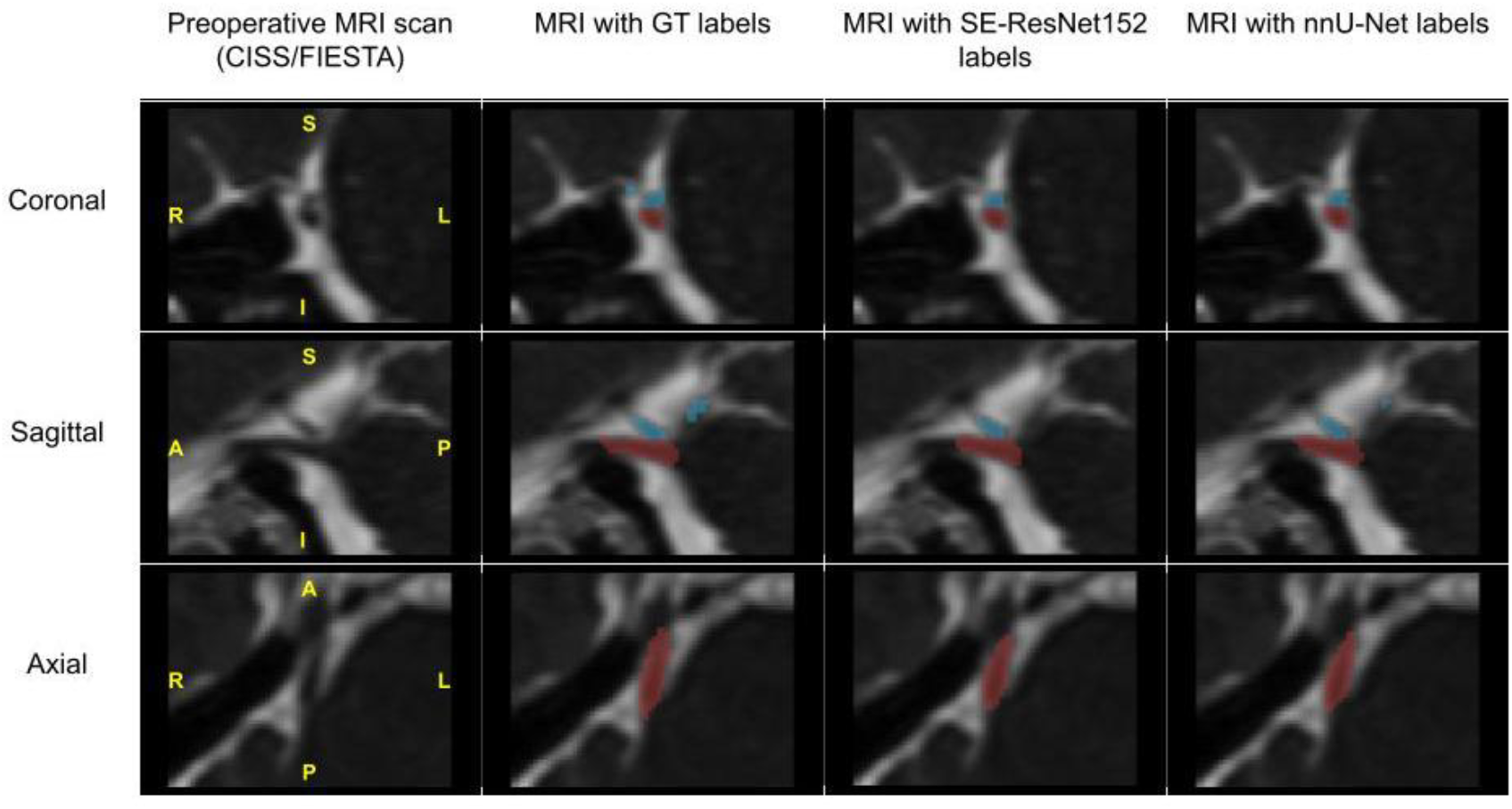
Coronal, sagittal, and axial slice orientations of a preoperative CISS/FIESTA MRI sequence with manual GT, SE-ResNet152-predicted, and nnU-Net-predicted labels of the trigeminal nerve (red) and nearby vasculature (blue).

**Figure 3.**
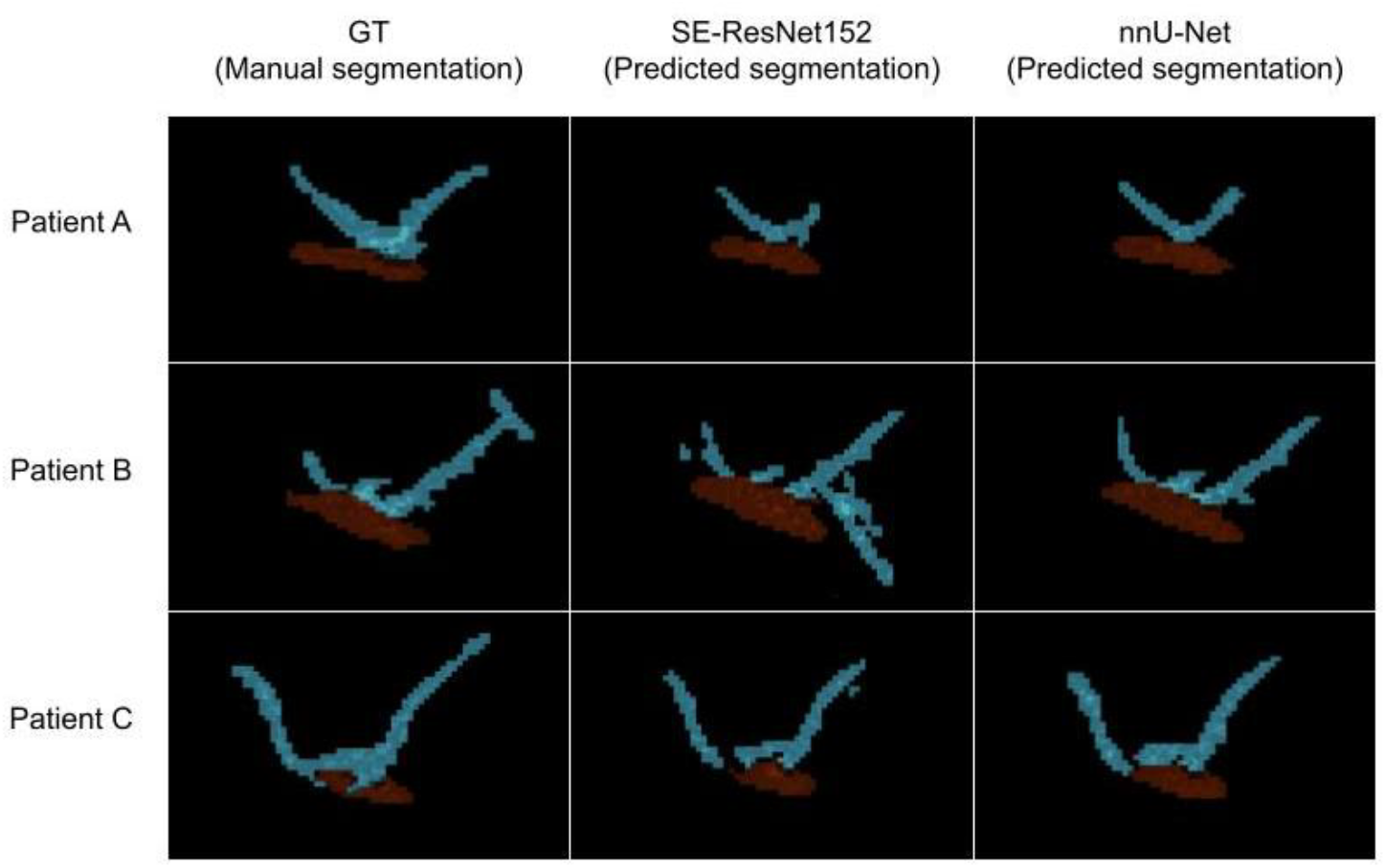
Three-dimensional visualizations of manually-generated (GT), SE-ResNet152-generated, and nnU-Net-generated segmentations of the trigeminal nerve (red) and nearby vasculature (blue).

### Segmentation post-processing

To suppress noise in the final segmentations, post-processing was performed to remove disconnected volumes of nerve or vessel totaling less than 10.4 mm^3^ (10 cubic voxels) in volume. The post-processing step also removed all but the largest contiguous structure labeled as nerve. Vessels close to (nearest point within 3 mm) or contacting the trigeminal nerve were kept for evaluation, as these are most likely to be the drivers of pathologic pain.

### Model evaluation

Each segmentation prediction in the testing dataset was evaluated against its corresponding GT segmentation with an F1 (also referred to as Dice) score, intersection-over-union (IoU) score, and a paired comparison of segmentation-calculated surface areas of NVC as described in our previous work.^15,19^ The F1 and IoU scores are defined as 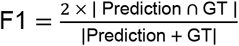 and 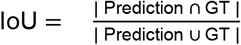, with “Prediction” and “GT” referring to the label volumes in the predicted segmentations and GT segmentations, respectively.^20^ The F1 score is the ratio of overlap volume to total volume, and the IoU score is the ratio of overlap volume to the volume of union. For both metrics, scores range from 0-1, with a score of 0 indicating no overlap between the two 3D models and a score of 1 indicating complete overlap. The F1 and IoU scores were calculated for nerve-only and vessel-only subsets of the full segmentation and also averaged between the labels for an overall score. The performances of the nnU-Net and SE-ResNet152 models according to F1 and IoU scores were compared, and the higher-fidelity model was selected for downstream analysis.

The sensitivity 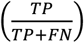 and specificity 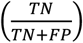 of the selected model to detect NVC in the testing dataset were calculated, with NVC defined as a minimum of one contact point between nerve and vessel.^19^ A contact point was further defined as a continuous region of shared voxel faces between nerve and vessel labels totaling more than 1.33 mm^2^ (6 square voxels) in area. This threshold was implemented to account for inherent limitations in MRI resolution, as distinguishing true NVC from proximity of nerve and vessel can be ambiguous for small interactions. The specific threshold value of 1.33 mm^2^ was chosen to maximize the true positive rate and minimize the false positive rate for NVC detection per receiver operating characteristic analysis (**Supplementary Figure 1**). Lastly, the surface area of NVC was calculated for each segmentation prediction and its corresponding GT segmentation. Surface area of contact was defined as the summed area of all voxel faces shared between nerve and vessel labels.

### Inference dataset

To evaluate the effectiveness of using nnU-Net segmentations to predict post-MVD outcomes, we applied the trained nnU-Net model to obtain nerve and vessel segmentations from the inference dataset MRIs, which were not used in the training and testing process of nnU-Net.

The same post-processing steps as those for training and testing were applied to these segmentations. The presence of NVC and surface area of NVC were derived from these segmentations, and these two datapoints were subsequently correlated with post-MVD patient pain recurrence. Pain recurrence was defined as a score of 3 or higher on a modified Barrow Neurologic Institute pain scale at final available follow-up or phone call.^4^

### Statistical analysis

Continuous variables are reported as means and standard deviations unless otherwise stated. Categorical variables are reported as counts. A paired Wilcoxon signed-rank test was conducted to test the null hypothesis that there is no significant difference between nnU-Net-derived and GT-derived calculations for surface area of NVC. Surface area of NVC was compared between patients with and without pain recurrence via a Wilcoxon rank-sum test.

Lastly, presence of NVC and surface area of NVC were correlated with postoperative pain-free survival using univariable Cox proportional hazards models. Significance level for all statistical tests was set at *α=0*.*05*. All statistical analysis was performed in R version 4.3.1 (The R Foundation).

## Results

A total of 332 patients underwent MVD at our institution from January 2019 to December 2022, of whom 283 (85.2%) were included in the study. Ten (3.0%) patients were excluded for prior MVD, 20 (6.0%) patients were excluded for having MRI quality issues (corrupted imaging file, presence of motion artifacts, insufficient capture of the trigeminal nerve), and 14 (4.2%) patients were excluded for having no available CISS/FIESTA MRI sequence. Within the inference dataset used for correlation with post-MVD clinical outcomes, 5 (1.5%) patients were excluded for having multiple sclerosis. For this overall cohort (n=283), the mean (±SD) age of the overall cohort was 57.2±14.3 years, and 109 (38.5%) of all patients were male; 221 (78.1%) and 62 (21.9%) patients presented with type I and type II TN pain, respectively; 173 (61.1%) patients presented with right-sided pain.

Randomly split across the 2020-2022 patient cohort (n=183), the training dataset consisted of 151 (82.5%) patients and the testing dataset consisted of 32 patients (17.5%). For the training and testing datasets, the mean (±SD) age was 56.7±14.2 years, and 76 (41.5%) patients were male; 136 (74.3%) and 47 (25.7%) patients presented with type I and type II TN pain, respectively; 107 (58.5%) patients presented with right-sided pain. From these 183 patients, 366 GT segmentations were manually created: 302 (82.5%) served to train the nnU-Net and SE-ResNet152 models, and 64 (17.5%) were used to evaluate their corresponding model predictions. The F1 and IoU scores comparing nnU-Net predictions with GT segmentations for the testing dataset were 0.797±0.011 and 0.714±0.011, respectively, while the F1 and IoU scores for SE-ResNet152 were 0.770±0.011 and 0.686±0.011, respectively (**Table 1**).

**Table 1.** Performance of nnU-Net and SE-ResNet152 on classifying nerve and nearby vasculature in preoperative TN imaging.

| Model | Overall |  | Nerve-only |  | Vessel-only |  |
| --- | --- | --- | --- | --- | --- | --- |
|  | F1 | IoU | F1 | IoU | F1 | IoU |
| nnU-Net (mean ± SD) | 0.797 ±0.011 | 0.714 ±0.011 | 0.828 ±0.010 | 0.713 ±0.013 | 0.575 ±0.010 | 0.444 ±0.030 |
| SE-ResNet152 (mean ± SD) | 0.770 ±0.011 | 0.686 ±0.011 | 0.808 ±0.010 | 0.684 ±0.012 | 0.517 ±0.034 | 0.397 ±0.034 |

nnU-Net was the higher-performing model per F1 and IoU analysis, indicating that predicted segmentations generated by nnU-Net were more similar to the GT segmentations than those by SE-ResNet152. Accordingly, the nnU-Net model was selected for subsequent analysis with clinical outcomes. nnU-Net correctly identified 42 of the 46 segmentations with NVC and 12 of the 18 segmentations without NVC, yielding a sensitivity of 91.3% and specificity of 66.7% when compared to GT. Furthermore, surface area of NVC calculated from nnU-Net segmentations was statistically similar with that from GT segmentations (nnU-Net: median [IQR] 6.53 [9.00] mm^2^; GT: 7.84 [12.5] mm^2^; *p=0*.*219*).

The inference dataset consisted of the 2019 patient cohort (n=100): the mean (±SD) age was 58.0±14.6 years and 33 (33.0%) of all patients were male; 85 (85.0%) and 15 (15.0%) patients presented with type I and type II TN pain, respectively; 66 (66.0%) patients presented with right-sided pain. The trained nnU-Net model generated a segmentation on the side ipsilateral with symptomatic pain for all 100 patients in the inference dataset, among whom 86 (86.0%) had NVC present. These 100 patients had a mean (±SD) final follow-up period of 21.4±21.7 months, and 30 (30.0%) patients experienced pain recurrence. Notably, patients without pain recurrence had a median (IQR) NVC surface area of 8.06 (9.94) mm^2^, which was significantly higher than the 4.31 (5.96) mm^2^ for patients with pain recurrence (*p=0*.*008*) (**Figure 4**). In the univariable Cox proportional hazards models, surface area of NVC (hazards ratio [HR] 0.914 per mm^2^, 95% confidence interval [CI] 0.848–0.985, *p=0*.*019*) and presence of NVC (HR 0.369 relative to absent NVC, 95% CI 0.156–0.876, *p=0*.*024*) were both significantly associated with decreased risks of pain recurrence (**Table 2**).

**Table 2.**
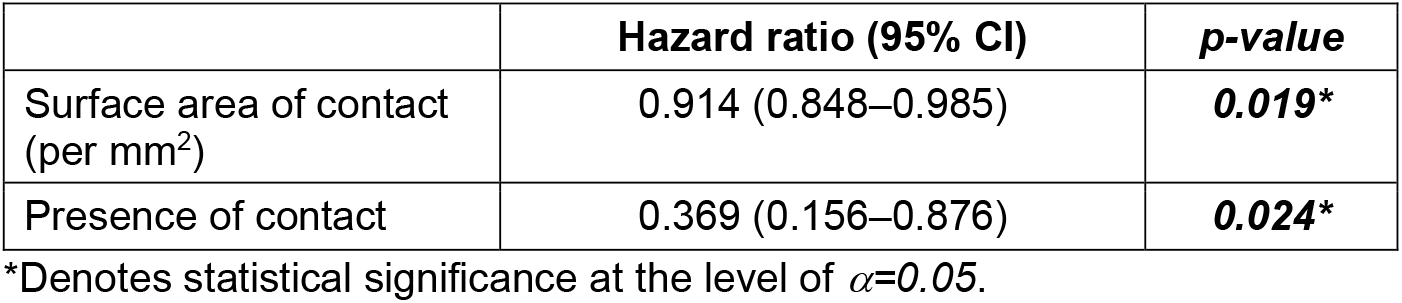
Univariable Cox proportional hazards models assessing the association between risk of pain recurrence and nnU-Net-derived surface area and contact presence metrics.

**Figure 4.**
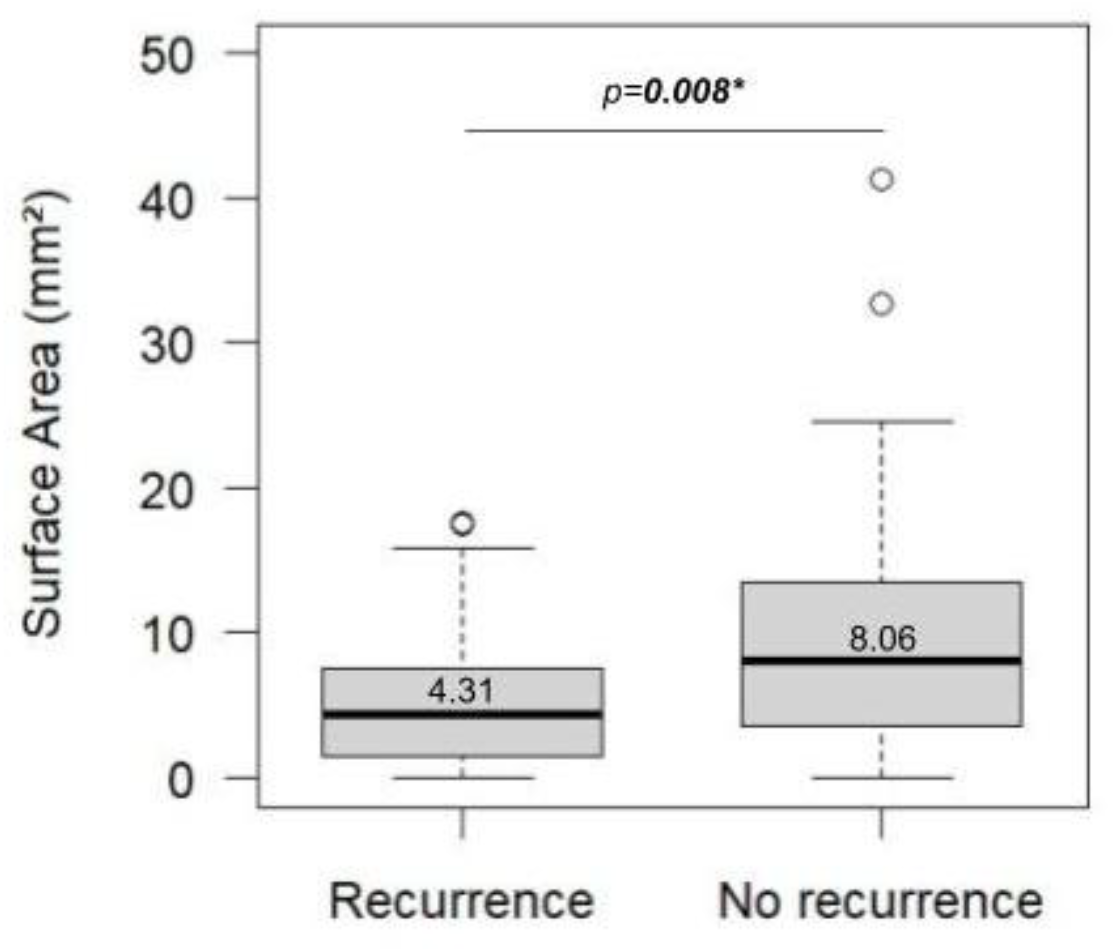
Distribution of nnU-Net-derived NVC surface area metric by post-MVD pain recurrence outcome (patients with recurrence, n=30; patients without recurrence, n=70).

## Discussion

Deep learning-generated segmentations possess several advantages over traditional MRI evaluation. Currently, there is a lack of standardized metrics to quantify NVC in clinical imaging reads—existing descriptors of nerve-vessel interactions such as “contacting,” “abutting,” “coursing along,” and “compressing” do not have specific definitions and accordingly remain qualitative.^5–7^ Segmentation analysis can yield more precise, quantitative measurements, such as surface area of NVC. However, inter-reader variability may be present in manually-created segmentations. Deep learning models are not subject to this variability as, once model parameters are fixed after training, they generate segmentations by applying the same algorithm to any input patient data. Moreover, its near-instantaneous timescale for inference offers another advantage over time-intensive manual segmentations and clinical reads.^8,13^

In this study, we found that the nnU-Net deep learning method can generate accurate segmentations of the trigeminal nerve and surrounding vasculature. Compared to a SE-ResNet-based model, nnU-Net featured higher F1 and IoU scores and was thus selected for analysis with clinical outcomes. Furthermore, nnU-Net is advantageous given its accessibility and availability as an open-source model, allowing deep learning to be implemented by clinicians regardless of their training background.

Importantly, we found that segmentation-derived metrics from nnU-Net were correlated with clinical post-MVD outcomes. Higher surface area and presence of NVC were each associated with a decreased risk of pain recurrence. Similarly, surface area of NVC was significantly higher in patients without pain recurrence than in patients with pain recurrence. These findings may be explained by the hypothesis that MVD most benefits TN patients who have greater magnitudes of NVC, a pathophysiology that can be successfully resolved by physical separation of nerve and vessel via MVD. Contrarily, patients with less significant NVC may have pain for which nerve-vessel interactions are only partially responsible.^21,22^ Accordingly, these patients may exhibit a weaker response to MVD and thus experience higher rates of pain recurrence.^21,23^ The correlation between NVC surface area and TN outcomes suggests that nnU-Net-generated segmentations can offer novel insights into the relationship between NVC severity and clinical TN, potentially enhancing evaluation and counseling for patients with TN.

Recently, Ishiwada et al. quantified several anatomical features of the trigeminal nerve in patients with TN and found that differences in centerline length, curvature, and cross-sectional area were present in the trigeminal nerve ipsilateral with symptomatic TN pain.^24^ Our work supports their hypothesis that quantitative anatomical features will be useful for disease evaluation, and further builds upon these efforts in three key ways. First, by using deep learning-generated segmentations instead of manual analysis, the process of labeling anatomy on MRI is more standardized and efficient. Second, our approach segmented the surrounding vasculature in addition to the trigeminal nerve itself, enabling us to analyze not only nerve anatomy but also vessel anatomy and features of nerve-vessel interaction. Accordingly, the metric used in our study, surface area of NVC, is only quantifiable using our methods. Third, we directly demonstrated the clinical relevance of quantitative metrics by investigating their association with post-MVD TN outcomes.

Quantitative metrics provided by our nnU-Net model can improve patient selection for surgical TN intervention. Surface area of NVC adds precise information not readily apparent from MRI reads alone, and has the potential to help clinicians identify TN patients who are most likely to experience sustained pain relief following MVD. Future work will study additional segmentation-based metrics, such as location of NVC along the trigeminal nerve and structural nerve changes secondary to compression.^11,25,26^ Combined with the readily generalizable ability of our model to generate a segmentation for any patient with a high-resolution preoperative brain MRI, this nnU-Net model can serve as the foundation for the development of an increasingly standardized and clinically-informative evaluation of NVC for all patients with TN.

### Limitations

The single-center nature of this study indicates that our findings may not be readily generalizable to other medical centers and other MRI machines. Additionally, our institution utilizes a preoperative CISS/FIESTA MRI sequence tailored specifically towards diagnosis of TN, which may differ from those used at other institutions. Furthermore, given the relative rarity of TN due to arteriovenous malformations, tumors, and other structural compression, such patients were not included in the model training and evaluation.

## Conclusion

nnU-Net can generate high-fidelity segmentations of the trigeminal nerve region to characterize the degree of NVC present in TN patients undergoing evaluation for MVD. Derived from nnU-Net-generated segmentations, higher surface area of NVC and presence of NVC are metrics that correlate clinically with decreased likelihood of pain recurrence following MVD. The nnU-Net model can help physicians better understand the relationship between NVC and postoperative outcomes, while clinically enhancing the process of treatment selection and counseling for patients with TN.

**Supplementary Figure 1.**
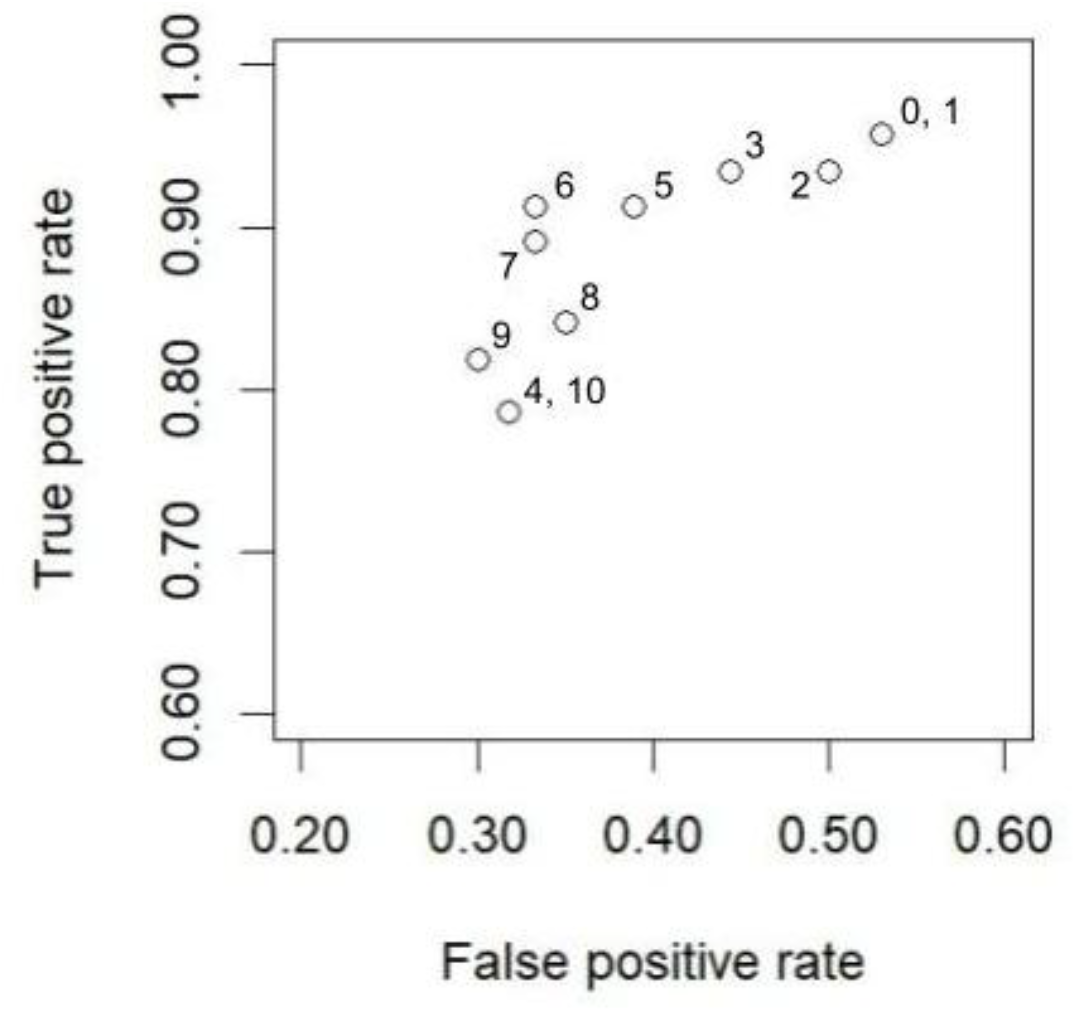
Receiving operating characteristic analysis showing false positive and true positive rates for NVC contact thresholds (square voxels).

## Data Availability

All data produced in the present study are available upon reasonable request to the authors.

